# The Power of Open Health Data: Impact, Representation, and Knowledge Diffusion

**DOI:** 10.64898/2026.03.20.26348933

**Authors:** Rahul Gorijavolu, Miguel Ángel Armengol de la Hoz, Catherine Bielick, Sebastian Cajas, Marie-Laure Charpignon, Aya El Mir, Judy W. Gichoya, Hyunjung Gloria Kwak, Kaushik Madapati, Heather Mattie, Lucas McCullum, Rogers Mwavu, Vishnu Nair, Luis Filipe Nakayama, Josephine Nanyonjo, Lama Nazer, Milit S. Patel, Christopher M. Sauer, Leo A. Celi

## Abstract

**Background:** Open health data repositories receive billions in public funding, yet no systematic framework exists to evaluate their downstream scholarly impact, the composition of the research communities they cultivate, or the breadth of disciplines they reach. We introduce a two-degree citation methodology to quantify knowledge diffusion from open data, normalized by funding, and apply it to four major health data repositories.

**Methods:** Using the OpenAlex bibliometric database (January–February 2026), we identified all first-degree citing publications (*n* = 30,049) and their second-degree citing publications (*n* = 485,396), defined as papers citing those first-degree publications, for MIMIC (versions I– IV; retrospective EHR data; $14.4M), UK Biobank (prospective cohort with genomics; $525.5M), OpenSAFELY (federated EHR platform; $53.7M), and All of Us (prospective national cohort with biobanking and community engagement; $2,160M). We extracted author demographics (gender via Genderize.io, institutional country income via World Bank 2024 classifications) and research topics. Chi-square tests with odds ratios assessed demographic differences across repositories.

**Results:** Funding-normalized first-degree papers per $1M ranged from 689 (MIMIC) to 1 (All of Us), though these figures reflect total program investment, which included community engagement and biobanking for prospective cohorts in addition to data-curation costs. The citation amplification ratio was consistent across these four repositories (9.3–11.5×). Author demographics differed significantly (*p <* 0.001): LMIC authorship ranged from 41.8% (MIMIC) to 4.3% (All of Us), while female authorship showed the opposite pattern, lowest for MIMIC (31.8%) and highest for All of Us (43.2%). Female authors were consistently underrepresented in senior (last-author) compared with first-author positions across all repositories. Differences in scope, design, and what funding covers limit direct comparisons.

**Conclusions:** Open health data generates a consistent ∼10× indirect citation amplification beyond its direct users, a ratio that held across repositories spanning over two orders of magnitude in funding. The large differences in funding-normalized output partly reflect structural differences between retrospective databases and prospective cohorts. Low-cost access combined with intentional community building attracted globally diverse research communities with LMIC investigators in intellectual leadership positions, while a persistent gender gap in senior authorship across all repositories reflects disciplinary and structural inequities that data access policies alone cannot address. Future evaluations of open data investments should examine who is producing research, from where, in what positions, and whether their participation translates into locally relevant knowledge production.

**Take-home message:** Open health data shows a consistent ∼10× citation amplification ratio beyond direct users. Low-barrier access and active community-building are associated with globally diverse research communities, including LMIC researchers in leadership positions, yet representational equity in authorship does not guarantee locally relevant knowledge production. A persistent gender gap in senior authorship across all repositories reflects disciplinary and structural inequities that data access policies alone cannot address.

## Introduction

Open data sharing has become a cornerstone of biomedical research policy. Major funding agencies including the U.S. National Institutes of Health and the Wellcome Trust now mandate that publicly funded datasets be made accessible, reflecting a growing consensus that open data accelerates scientific discovery [1–4]. This commitment has translated into substantial public investment: in health sciences alone, programs such as the All of Us Research Program ($2.16 billion), UK Biobank ($525.5 million), and the MIMIC critical care database ($14.4 million) span orders of magnitude in funding yet share the common goal of enabling secondary research [5–7].

Despite these investments, there is no systematic framework for evaluating the downstream scholarly impact of open health data repositories. Existing bibliometric evaluations typically count publications or first-degree citations, referencing papers that directly use a dataset, but fail to capture the broader knowledge diffusion that occurs when those papers are themselves cited by others [8, 9]. This indirect effect is critical because the value of open data extends well beyond its immediate users: a clinical prediction model built on MIMIC may be cited by implementation science researchers, health economists, or computer scientists who never access the original data.

Equally important is the question of who comprises the research communities that open data cultivates. Global disparities in scientific publishing are well documented, and open data has been proposed as a mechanism for leveling the playing field by providing researchers in low- and middle-income countries (LMICs) access to datasets they could not independently generate [10, 11]. Yet whether this promise is being realized, and whether open data attracts diverse researchers into positions of intellectual leadership, not merely participation, remains an empirical question. Understanding who does the research, where they are located, and what positions they hold in the authorship hierarchy is as important as counting how many papers a dataset generates. This requires distinguishing between representational equity, defined as the presence and positioning of diverse researchers in authorship, and transformative equity, which would encompass whether open data reshapes research agendas, builds local capacity, or produces locally relevant knowledge [12, 13].

In this study, we introduce a two-degree citation methodology that traces both direct citations (first-degree: papers that use a dataset) and indirect citations (second-degree: papers citing those first-degree papers) to quantify knowledge diffusion from open data, normalized by cumulative program funding. We apply this framework to four major open health data repositories spanning different designs, maturity stages, and funding scales: MIMIC (versions I–IV; retrospective electronic health records), UK Biobank (a prospective population cohort with genomic data), OpenSAFELY (a federated analytics platform for primary care records), and All of Us (a prospective national cohort emphasizing participant diversity) [5–7, 14–17].

Our objectives are: (1) to quantify funding-normalized scholarly output at both degrees of citation, (2) to characterize the demographics of the research communities each repository has cultivated, including gender representation, geographic diversity, and authorship position, and (3) to examine how equity patterns vary across repositories that serve fundamentally different research communities.

## Methods

### Study Design

This cross-sectional bibliometric analysis included publications from each repository’s first public data release through December 31, 2025.

### Data Repositories

We selected four large open health data repositories representing distinct models of data sharing in biomedicine (Table 1).

**Table 1:**
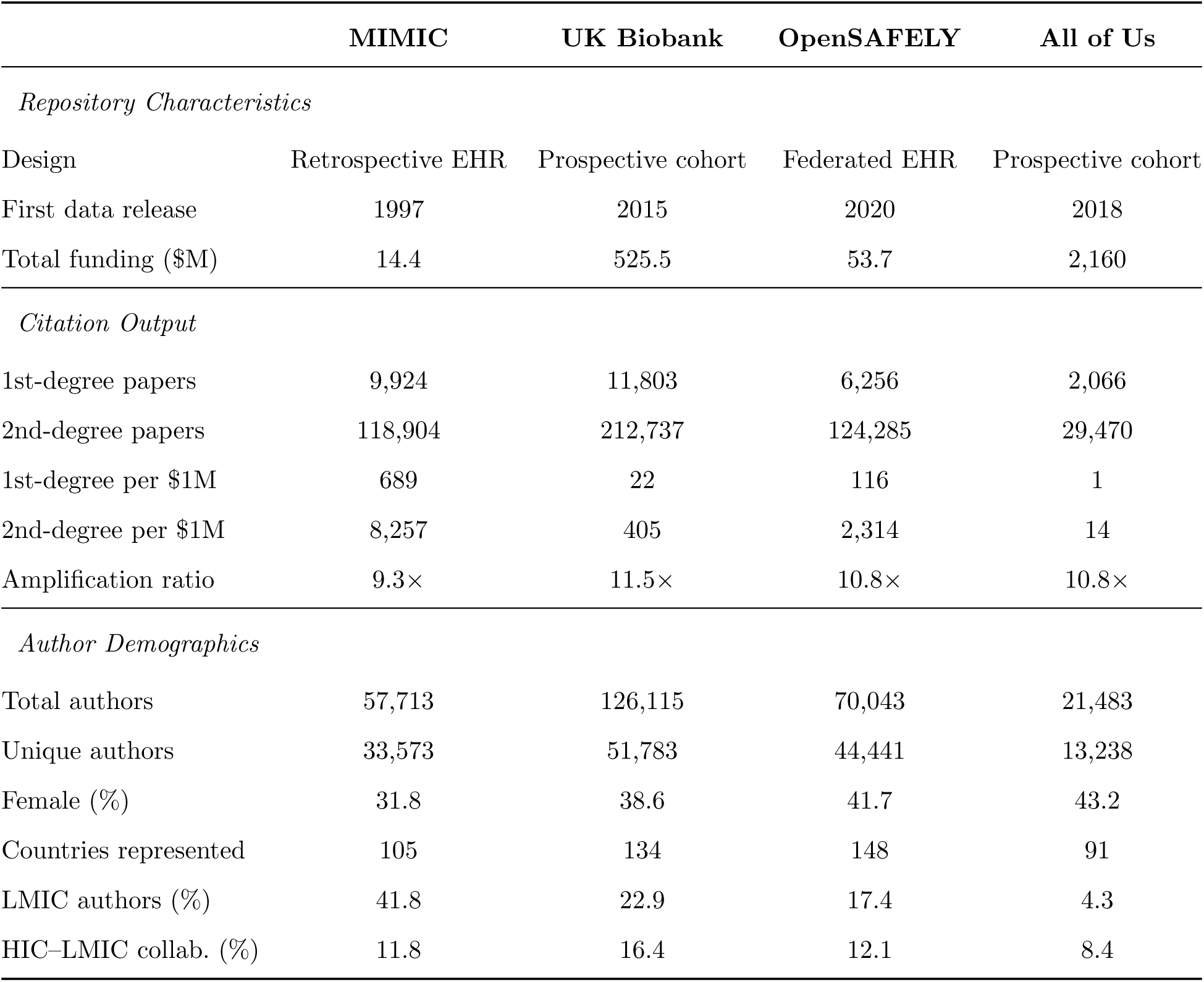
Repository characteristics, citation output, and author demographics. Funding figures represent total program investment to date. First-degree papers are publications directly citing a repository’s primary works; second-degree papers cite those first-degree papers. Author demographics are computed across all first-degree publications. Gender was inferred via Genderize.io (identification rate *>*98% for all repositories). Country income classification follows World Bank 2024 definitions.

MIMIC (Medical Information Mart for Intensive Care) is a family of freely accessible critical care databases originating from Beth Israel Deaconess Medical Center. MIMIC-I was first released in 1997, with subsequent versions (II, III, IV) expanding in scope and detail [7, 14, 18]. MIMIC is a retrospective database built from routinely collected EHR data, with a patient population representing a predominantly insured, urban U.S. population in Boston, MA. Its total estimated funding is $14.4 million.

UK Biobank is a prospective cohort study of approximately 500,000 participants aged 40–69, with data available for approved research since 2015 [6]. The resource includes genotyping, imaging, and longitudinal health outcomes, with cumulative funding of approximately $525.5 million [15].

OpenSAFELY is a secure, federated analytics platform providing access to de-identified primary care records of approximately 58 million patients in England [16]. Launched during the COVID-19 pandemic in 2020, it has received approximately $53.7 million in grant funding [19].

All of Us is a longitudinal research program of the U.S. National Institutes of Health aiming to enroll one million or more participants reflecting the diversity of the United States, with data first available to researchers in 2018 [5]. Its funding encompasses participant recruitment, biobanking, genomic sequencing, return of results, and community engagement infrastructure, totaling approximately $2.16 billion [17].

Funding estimates were compiled from publicly available grant databases, published reports, and institutional websites.

### Citation Data Collection

Using the OpenAlex bibliometric database, we identified all publications citing each repository’s primary works (first-degree data retrieved January 2026; second-degree data retrieved February 2026) [20]. For MIMIC, we pooled citations across all four versions (I–IV); second-degree papers were deduplicated by OpenAlex work identifier. These citing publications constitute the first-degree corpus. We then retrieved all publications citing these first-degree papers, forming the second-degree corpus. In total, we identified 30,049 first-degree and 485,396 second-degree publications.

### Author Demographics

Author gender was determined from first names using the Genderize.io API, a validated tool that returns probabilistic gender assignments based on a global name–gender database [21]. The identification rate exceeded 98% across all repositories.

Institutional country was determined from author affiliation metadata in OpenAlex. Countries were classified as high-income (HIC) or low- and middle-income (LMIC) using the World Bank 2024 income classification [22]. HIC–LMIC collaboration was defined as any publication with authors from both income groups.

We analyzed authorship positions separately: first authors (typically the primary researcher), last authors (typically the senior or supervising investigator), and middle authors. This distinction is important because first- and last-author positions carry different implications for intellectual leadership, career advancement, and mentorship roles within research teams.

Name-based gender inference has known limitations: it performs systematically worse for East African, South and Southeast Asian, and Arabic names, conflates gender identity with name-associated gender, and cannot capture non-binary identities. [23] Given MIMIC’s LMIC authorship concentrates in regions where Genderize.io performs less reliably, gender estimates for LMIC authors may be less accurate than the identification rate (*>*98%) suggests. Identification rates by World Bank region are reported in the Supplementary Materials (Table S4). Institutional affiliation may not reflect nationality or career trajectory. These proxies are standard in large-scale bibliometric analyses but should be interpreted as approximations.

### Disciplinary Analysis

OpenAlex assigns each publication a hierarchical topic classification. We extracted the primary field assignment for each first-degree publication and computed field proportions by repository and year. Full disciplinary diffusion results are reported in the Supplementary Materials (Figure S1).

### Statistical Analysis

Differences in author demographics across repositories were assessed using Pearson’s chi-square tests, with Craméer’s *V* as the effect size. Pairwise comparisons between MIMIC and each other repository used chi-square tests with Bonferroni correction (*n* = 3). Odds ratios (OR) with 95% confidence intervals were computed for all 2 × 2 comparisons. The significance level was set at *α* = 0.05.

Funding-normalized metrics were computed by dividing raw counts by total program funding in millions of U.S. dollars. The citation amplification ratio was defined as the ratio of total second-degree citation mass (sum of all citations received by second-degree papers) to total first-degree citation mass (sum of all citations received by first-degree papers) for each repository. All analyses were conducted using R (version 4.3) and Python (version 3.11). Analysis code is publicly available at https://github.com/24rahul/power-of-data-communities.

### Ethical Approval and Consent

Patients and the public were not involved in the design, conduct, or reporting of this bibliometric analysis. No human participants were involved, and no ethics approval was required.

## Results

### Citation Output and Funding-Normalized Impact

Table 1 summarizes the citation output and funding context for each repository. First-degree papers ranged from 2,066 (All of Us) to 11,803 (UK Biobank). When normalized by funding, the variation was far more pronounced: MIMIC generated 689 first-degree papers per $1 million, compared with 116 (OpenSAFELY), 22 (UK Biobank), and 1 (All of Us).

This pattern amplified at the second degree. MIMIC’s 118,904 second-degree papers translated to 8,257 per $1 million, versus 2,314 (OpenSAFELY), 405 (UK Biobank), and 14 (All of Us). The cumulative second-degree citation mass per $1 million was 141,752 for MIMIC and 177 for All of Us, an approximately 800-fold difference. Despite this wide variation in funding-normalized output, the citation amplification ratio was relatively consistent across all four repositories: 9.3× (MIMIC), 11.5× (UK Biobank), 10.8× (OpenSAFELY), and 10.8× (All of Us).

Figure 1 shows the cumulative growth in first-degree papers over time, and Figure 2 presents funding-normalized first- and second-degree counts side by side. Figure 3 visualizes the cumulative funding-normalized citation trajectory as an “iceberg” plot, with first-degree citations above and second-degree citations below, illustrating the persistent ∼10× amplification ratio across all repositories.

**Figure 1:**
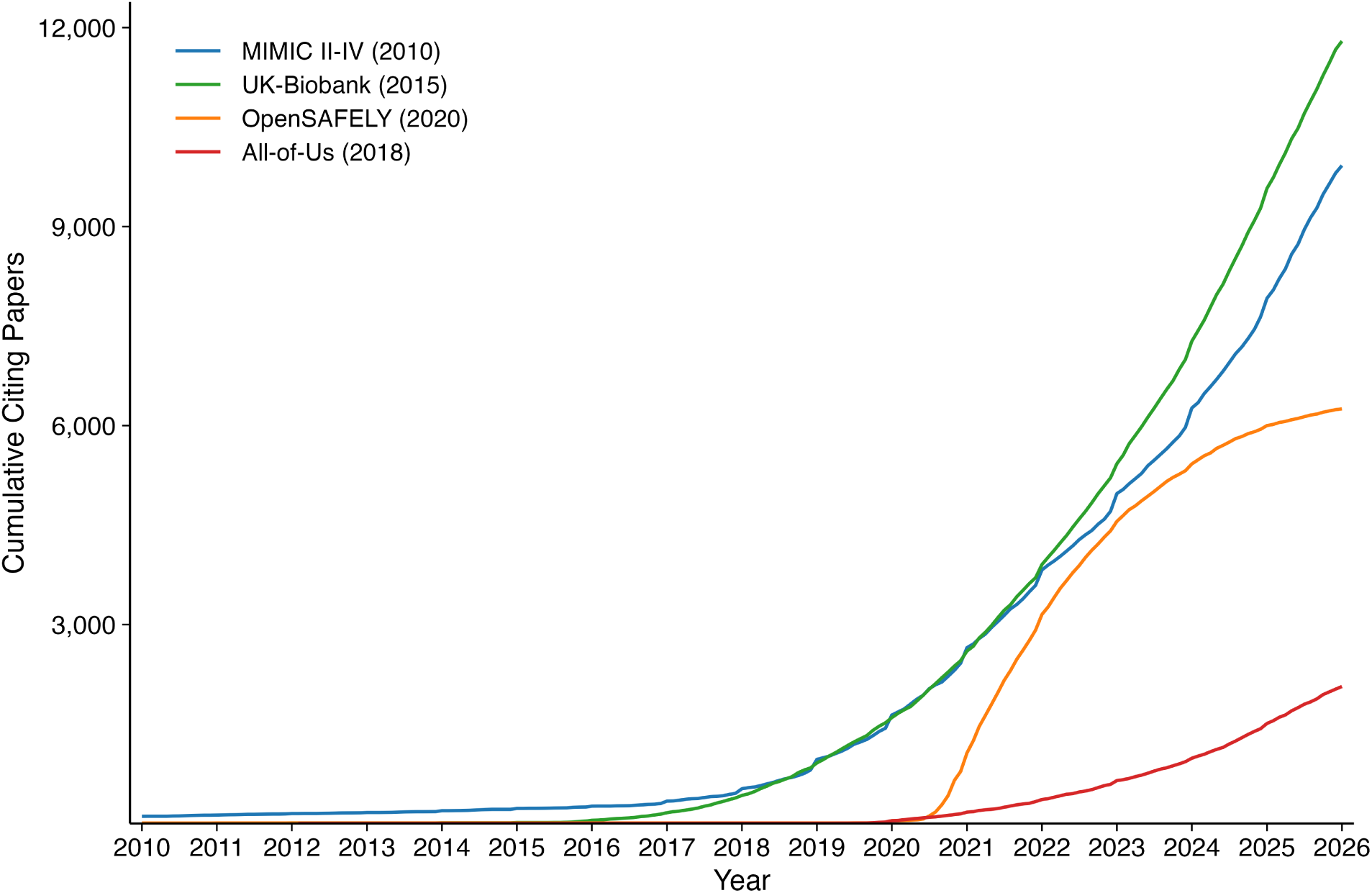
Cumulative first-degree publications over time for each repository (monthly resolution). Year of first data availability is shown in parentheses.

**Figure 2:**
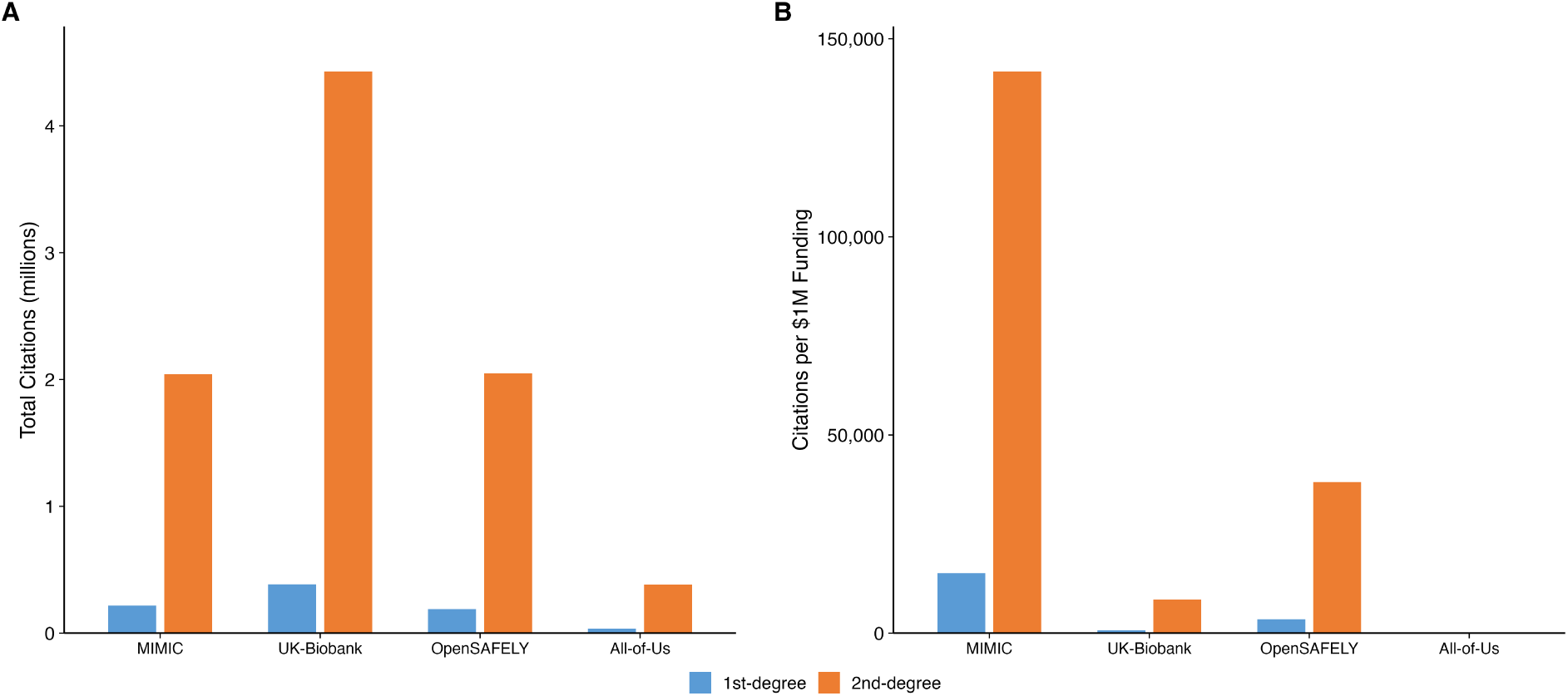
First-degree and second-degree citation counts. Panel A: absolute counts. Panel B: normalized per $1 million in funding.

**Figure 3:**
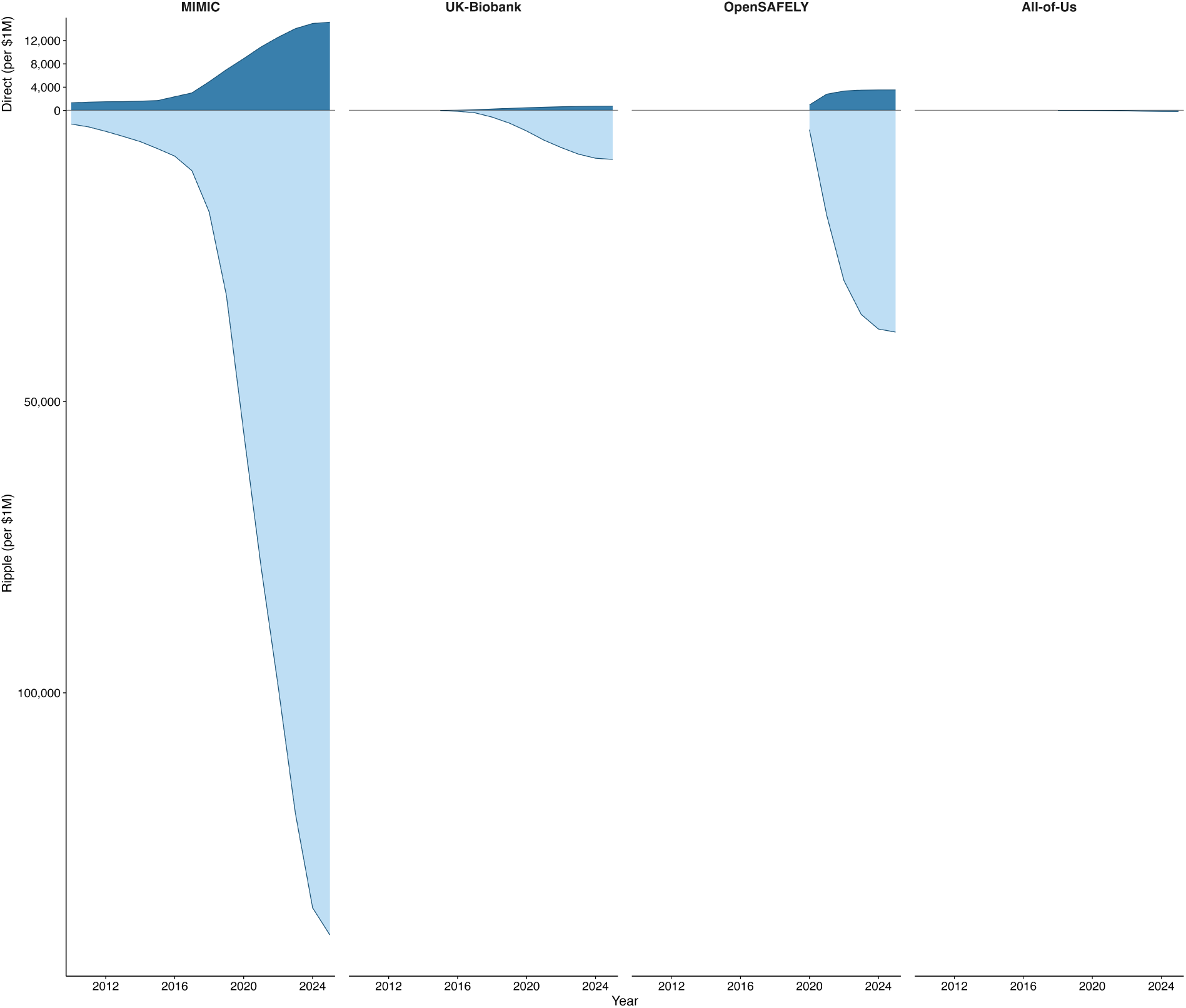
Cumulative funding-normalized citation trajectories. Panel A: first-degree (direct) citations per $1M. Panel B: second-degree (indirect) citations per $1M. Panel heights are proportional to maximum values. Note different y-axis scales.

### Author Demographics and Equity

Table 2 presents the full demographic profile of research communities across repositories, including authorship position analysis.

**Table 2:**
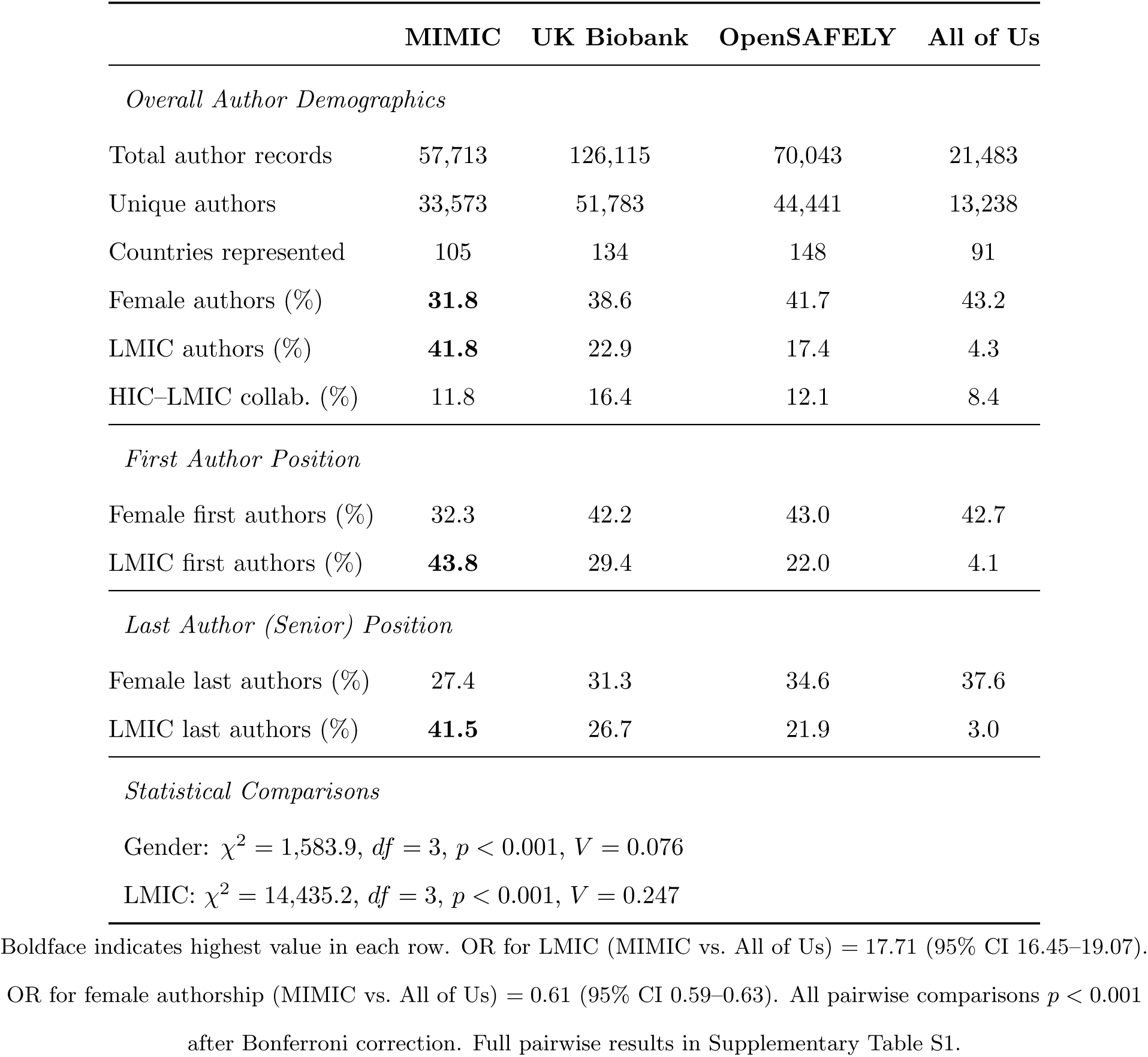
Author demographics and authorship position analysis across repositories. Gender was inferred via Genderize.io (identification rate *>*98% for all repositories). Country income classification follows World Bank 2024 definitions. HIC–LMIC collaboration indicates publications with authors from both income groups. First and last author positions are analyzed separately to distinguish primary researchers from senior investigators.

#### Geographic Diversity and LMIC Representation

Author demographics differed significantly across all four repositories for LMIC representation (*χ*^2^ = 14,435.2, *df* = 3, *p <* 0.001, *V* = 0.247). MIMIC had the highest proportion of LMIC-affiliated authors (41.8%), followed by UK Biobank (22.9%), OpenSAFELY (17.4%), and All of Us (4.3%). MIMIC’s odds of LMIC authorship were 17.7 times those of All of Us (OR = 17.71, 95% CI 16.45–19.07; *p <* 0.001).

Research citing these repositories originated from 91 (All of Us) to 148 (OpenSAFELY) countries. However, geographic breadth did not correspond with LMIC depth: OpenSAFELY drew authors from the most countries but had the second-lowest LMIC representation (17.4%). HIC– LMIC collaboration rates ranged from 8.4% (All of Us) to 16.4% (UK Biobank).

MIMIC’s high LMIC authorship extended to first- and last-author positions. LMIC researchers comprised 43.8% of first authors and 41.5% of last (senior) authors on MIMIC-citing papers, compared with 4.1% and 3.0% for All of Us, respectively. For UK Biobank, LMIC researchers held 29.4% of first-author and 26.7% of last-author positions; for OpenSAFELY, 22.0% and 21.9%.

#### Gender Representation

Gender distribution also differed significantly across repositories (*χ*^2^ = 1,583.9, *df* = 3, *p <* 0.001, *V* = 0.076). Female authorship was highest where LMIC authorship was lowest: All of Us had the highest female authorship (43.2%) and MIMIC the lowest (31.8%), with OpenSAFELY (41.7%) and UK Biobank (38.6%) between. MIMIC’s odds of female authorship were significantly lower than all comparators (OR = 0.61 vs. All of Us, 95% CI 0.59–0.63; OR = 0.65 vs. OpenSAFELY; OR = 0.74 vs. UK Biobank; all *p <* 0.001).

#### The Senior Authorship Gap

Across all four repositories, women were better represented in first-author positions than in last-author (senior) positions. The gap ranged from 4.9 percentage points (MIMIC: 32.3% first vs. 27.4% last; OR = 1.27, *p <* 0.001) to 10.9 percentage points (UK Biobank: 42.2% vs. 31.3%; OR = 1.61, *p <* 0.001). OpenSAFELY (43.0% vs. 34.6%; OR = 1.43, *p <* 0.001) and All of Us (42.7% vs. 37.6%; OR = 1.24, *p* = 0.001) showed intermediate gaps.

#### Intersectional Patterns

In three of the four repositories, the proportion of female authors was higher within the LMIC subgroup than within the HIC subgroup across most authorship positions (Table S2). Among MIMIC first authors, 36.4% of LMIC authors were female compared with 30.0% of HIC authors; among last authors, 31.9% versus 23.9%. Similar patterns held for UK Biobank (44.7% vs. 40.8% for first authors) and OpenSAFELY (46.2% vs. 42.6%). All of Us did not follow this pattern (40.0% vs. 43.1% for first authors), though the small LMIC sample sizes (*n* = 24–30 for first and last authors) limit interpretation.

### Disciplinary Analysis

Disciplinary analysis revealed that MIMIC is the only repository whose primary research field is computer science (43.3%) rather than medicine, reflecting its adoption as a machine learning benchmark. UK Biobank research is concentrated in biochemistry, genetics, and molecular biology, while OpenSAFELY and All of Us remain predominantly within clinical medicine (Figure S1).

## Discussion

### Principal Findings

We introduce a two-degree citation methodology to evaluate the scholarly impact and equity of four major open health data repositories. Three principal findings emerged: a consistent ∼10× citation amplification from direct to indirect citations, marked differences in the composition of each repository’s research community, with LMIC representation highest and female authorship lowest for MIMIC, and the reverse for All of Us, and a persistent gender gap in senior authorship across all repositories.

### The Research Communities Behind Open Data

The composition of the communities producing research on open data warrants as much attention as the volume of that research. MIMIC’s research ecosystem is distinctive: 41.8% LMIC authorship, researchers from 105 countries, and LMIC researchers holding first- and last-author positions at rates comparable to their overall representation. This pattern suggests a research community in which LMIC investigators exercise intellectual leadership, not merely participation.

However, this representational equity must be interpreted cautiously. MIMIC’s patient population originates from a single academic medical center in Boston, representing a predominantly insured, urban U.S. population. LMIC researchers building clinical prediction models or decision tools on this data are working with a population that may differ substantially from the patients they serve in disease burden, healthcare infrastructure, and care delivery patterns. Intellectual leadership in authorship hierarchies does not automatically translate into locally relevant knowledge. High LMIC authorship may also partly reflect computational benchmarking: computer science researchers developing machine learning methods using MIMIC as a convenient testbed rather than producing clinical decision tools for their local contexts.

Several structural features of MIMIC may contribute to its global reach. It is freely accessible with no financial barrier, requires only an online training course for credentialing, and operates on data small enough to be downloaded and analyzed on a personal computer. The extensive datathon program, which has engaged researchers at events in over 30 countries, provides hands-on mentorship and entry into collaborative networks. These features lower multiple barriers simultaneously: cost, infrastructure, and social capital.

By contrast, All of Us’s 4.3% LMIC authorship reflects the program’s design as a U.S.-focused cohort. Its data use agreements, institutional requirements, and cloud-based analysis platform may present barriers for international researchers. UK Biobank and OpenSAFELY show intermediate LMIC authorship (22.9% and 17.4%), consistent with their UK-centric data and governance structures.

High LMIC authorship on a dominant dataset does not, by itself, indicate equitable knowledge production. LMIC researchers using MIMIC produce research legible to Global North journals—a dynamic that benefits individual careers but does not necessarily challenge existing hierarchies of knowledge production. The more fundamental question is whether LMIC researchers’ engagement with MIMIC translates into investment in building open datasets within their own health systems [11]. Evaluating whether LMIC-generated datasets achieve comparable citation amplification would offer a more direct measure of progress toward transformative equity.

### The Gender Gap: Disciplinary Composition and Community Agency

MIMIC’s low female authorship (31.8%) appears paradoxical given its otherwise low-barrier access model. However, MIMIC is the only repository whose primary citing field is computer science (43.3% of papers; see Figure S1) rather than medicine, and the gender gap in computer science authorship is well documented [10, 24]. MIMIC’s female authorship rate is thus partly consistent with the demographics of its dominant citing discipline rather than a barrier intrinsic to the dataset.

That said, MIMIC is an actively cultivated ecosystem: its datathon program, credentialing community, and mentorship networks represent levers that could be used to improve gender balance, for instance through targeted recruitment of female participants in datathons or mentorship pairings. The persistent senior authorship gap, where women were 4.9 to 10.9 percentage points less represented in last-author than first-author positions across all four repositories, mirrors well-documented patterns in biomedicine [10, 25, 26]. This gap transcends individual datasets and reflects structural inequities in career advancement, mentorship, and access to independent funding, though repository communities with active mentorship programs have some agency to address it.

The intersectional analysis (Table S2) complicates a purely disciplinary explanation. In three of the four repositories, the proportion of female authors was higher within the LMIC subgroup than within the HIC subgroup, suggesting that the pathways through which researchers engage with open data may interact with gender representation in ways that merit further investigation.

### Funding Efficiency in Context

The approximately 800-fold difference in funding-normalized second-degree citations between MIMIC and All of Us requires contextualization. MIMIC retrospectively curates data that already exists in hospital systems; All of Us is building a prospective cohort from the ground up, with costs encompassing recruitment, biobanking, genomic sequencing, and community engagement; UK Biobank and OpenSAFELY occupy intermediate positions [5, 17]. The funding-normalized comparison therefore measures citation output per dollar of total program investment, not per dollar of data-curation cost. Programs that invest in participant engagement, biobanking, and community infrastructure produce value not captured by bibliometric analysis.

MIMIC’s high citation count also partly reflects its adoption as a machine learning benchmark, analogous to ImageNet in computer vision. When a dataset’s primary use shifts from clinical research to computational benchmarking, citation counts may reflect methodological convenience rather than clinical knowledge production.

These considerations cut in both directions. The funding-normalized gap overstates differences in data-curation efficiency because it includes non-data costs. At the same time, the community engagement, participant diversity, and biobanking investments made by programs like All of Us represent assets whose value is invisible to citation metrics. Future work could attempt to normalize by data-curation costs alone, which would substantially narrow the gap, or develop complementary metrics that capture these non-citation dimensions of impact.

### Data Provenance and Epistemic Justice

The equity dimensions of open data extend beyond who publishes to include whose data is used, collected under what conditions, and governed by whom. MIMIC’s adoption as a global benchmark means researchers worldwide are building clinical tools calibrated to a single U.S. hospital population, raising questions about whose health experiences are encoded as normative and whose are treated as exceptions [13, 27]. Current open data repositories operate on a spectrum from researcher-controlled (MIMIC) to community-partnered (All of Us), and emerging frameworks such as the CARE principles (Collective benefit, Authority to control, Responsibility, Ethics) offer models for more participatory governance [28]. Our bibliometric methodology cannot capture these governance dimensions, and developing metrics that measure community participation in dataset design, local capacity built, or policy influence would complement citation-based assessments.

### Strengths and Limitations

Strengths include the two-degree citation approach, analysis of over 515,000 publications, author-level demographic data, including position analysis, and a comparative framework spanning diverse repository designs.

Several limitations should be acknowledged. First, citation metrics capture scholarly activity but not necessarily clinical impact, policy influence, or community engagement. Second, funding figures are approximate and not strictly comparable; we could not decompose them into data-curation versus non-data costs. Third, gender inference from first names may misclassify individuals with culturally specific names and cannot capture non-binary identities, with Genderize.io performing less reliably for East African, South and Southeast Asian, and Arabic names (Table S4), which may differentially affect gender estimates for LMIC authors. Fourth, our two-degree methodology captures breadth but not depth of knowledge diffusion: a methods paper and a practice-changing clinical trial both generate second-degree citations but differ fundamentally in impact. Fifth, our analysis does not account for self-citation or citation network effects, which could inflate counts for more established datasets. Sixth, we did not disaggregate the citation amplification ratio by research type (e.g., clinical studies vs. computational benchmarking), which would reveal whether different research modes drive different ratios. We also did not distinguish positive, neutral, and negative citations, and therefore citation mass should not be interpreted as synonymous with scientific quality or translational value. Finally, the ∼10× amplification ratio is based on four repositories selected for diversity, not randomly sampled, and should be treated as an exploratory finding.

## Conclusions

Open health data generates scholarly impact well beyond its direct users, but impact is not equity. Low-barrier access can attract diverse researchers into leadership positions, yet representational equity in authorship does not guarantee locally relevant knowledge, and structural inequities like the senior authorship gender gap remain beyond the reach of data access policies alone. Future evaluations should move beyond citation metrics to examine whether participation translates into locally relevant knowledge production, local dataset creation, and capacity building.

## Data Availability

All data and analysis code used in this study are publicly available at https://github.com/
24rahul/power-of-data-communities with no restrictions.

https://github.com/24rahul/power-of-data-communities

## Acknowledgements

We thank the teams behind MIMIC, UK Biobank, OpenSAFELY, and the NIH All of Us Research Program for their commitment to making clinical data accessible for research. We acknowledge the broader data science and critical care research communities whose collaborative spirit and open-source contributions have fostered the ecosystem that made this analysis possible.

## Data Sharing Statement

All data and analysis code used in this study are publicly available at https://github.com/24rahul/power-of-data-communities with no restrictions. The repository includes: (1) complete citation datasets for all four repositories (MIMIC, UK Biobank, OpenSAFELY, All of Us) collected from OpenAlex as of February 2026; (2) funding data compiled from publicly available grant databases, published reports, and institutional websites; (3) all analysis scripts for calculating metrics and generating figures in R and Python; and (4) a comprehensive data dictionary describing all variables and their sources. These data are available immediately and indefinitely with publication under a Creative Commons Attribution 4.0 International (CC BY 4.0) license. No registration or approval is required to access or use these data and code. Researchers may freely download, modify, and redistribute the materials with appropriate attribution. Questions about the data or code should be directed to the corresponding author.

## Author Contributions (CRediT)

Conceptualization: RG, LAC. Data curation: RG, KM. Formal analysis: RG, KM. Visualization: RG, KM. Funding acquisition: CMS, LAC. Methodology: RG, KM, LM, VN, CMS. Project administration: CMS, LAC. Software: RG, KM. Supervision: LAC. Validation: RG, KM, VN. Writing – original draft: RG, MAAH, CB, SC, MLC, AEM, JWG, HGK, KM, HM, LM, RM, VN, LFN, JN, LN, MSP, CMS, LAC. Writing – review & editing: RG, KM, LM, VN.

## Ethical Approval

This study is a bibliometric analysis of publicly available citation data and did not involve human participants. No ethics approval was required.

## Supplementary Materials

**Figure S1:**
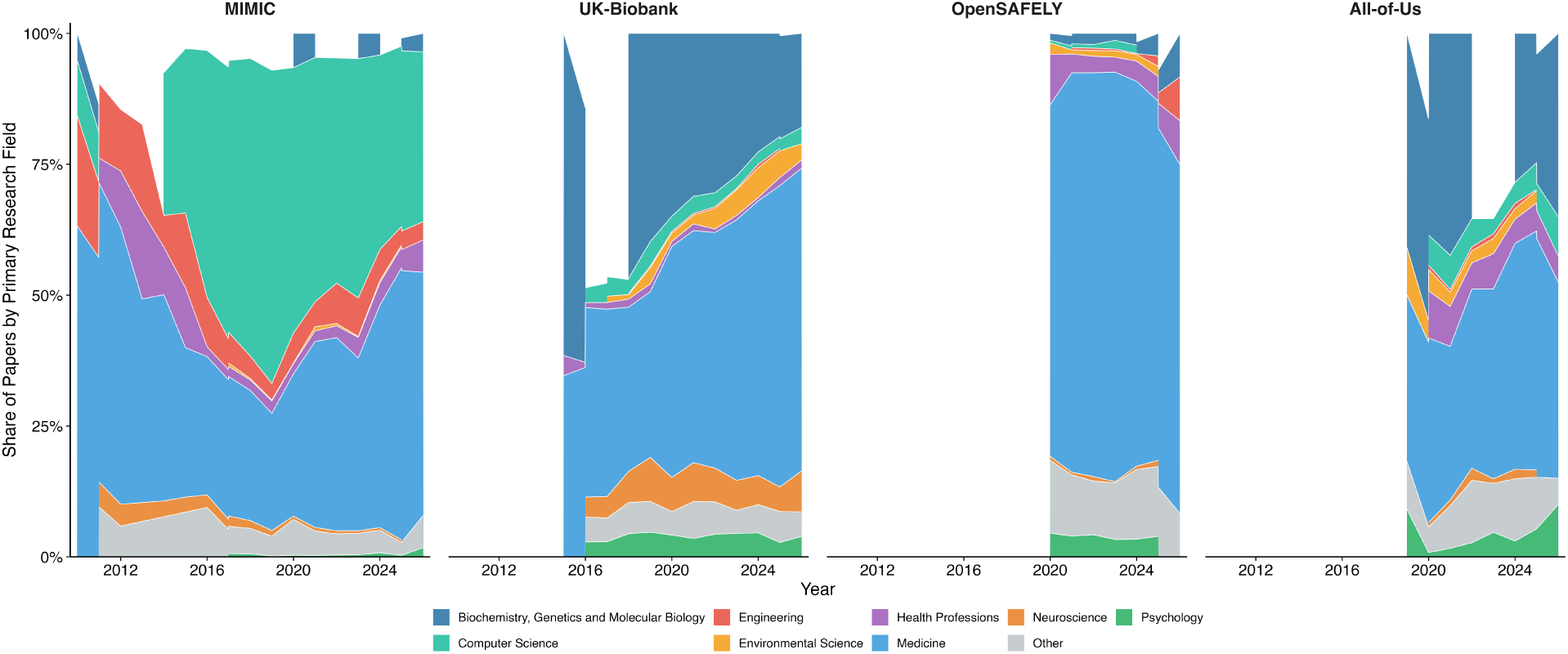
Disciplinary composition of first-degree publications over time by repository. Each color represents a research field as classified by OpenAlex. Only year–repository combinations with ≥10 papers are shown. MIMIC is the only repository whose primary citing field is computer science (43.3%) rather than medicine, reflecting its widespread adoption as a machine learning benchmark. UK Biobank is dominated by biochemistry, genetics, and molecular biology. OpenSAFELY and All of Us remain concentrated within medicine.

**Figure S2:**
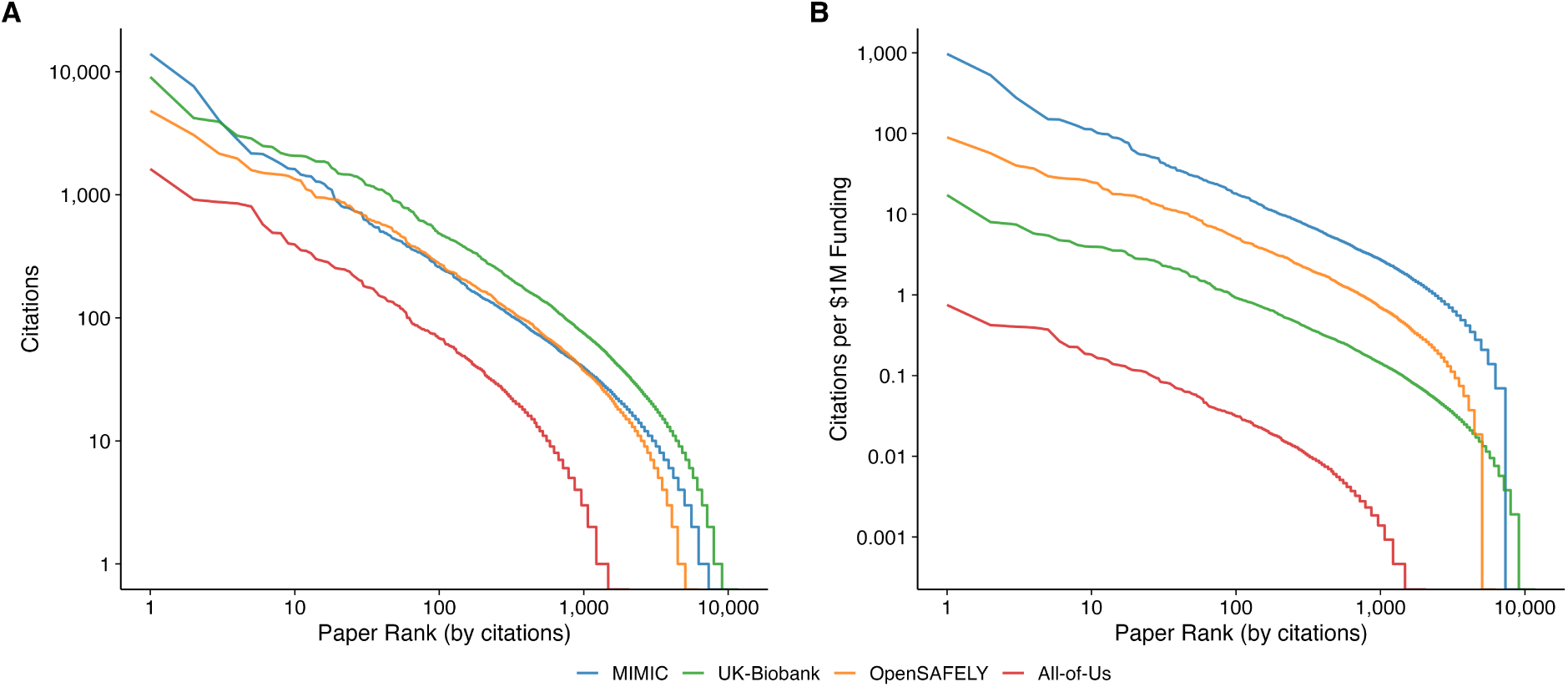
Citation distributions for first-degree publications. Panel A: Raw citation counts by paper rank (log-log scale). Panel B: Funding-normalized citations per $1M by paper rank (log-log scale). MIMIC papers are more highly cited per dollar of funding across the full rank distribution.

**Figure S3:**
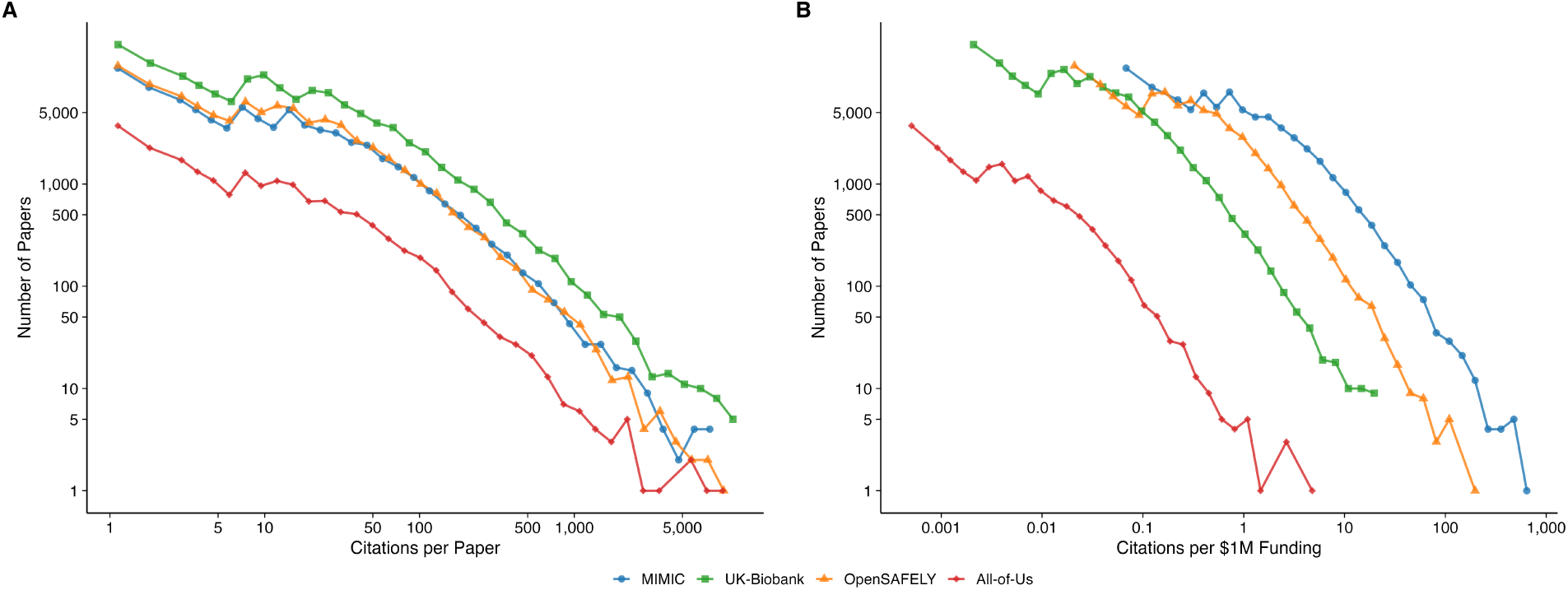
Citation distributions for second-degree publications. Panel A: Raw citation distribution (number of papers at each citation count). Panel B: Funding-normalized citation distribution. The distributions are broadly similar across repositories, consistent with the ∼10× amplification constant.

**Table S1:**
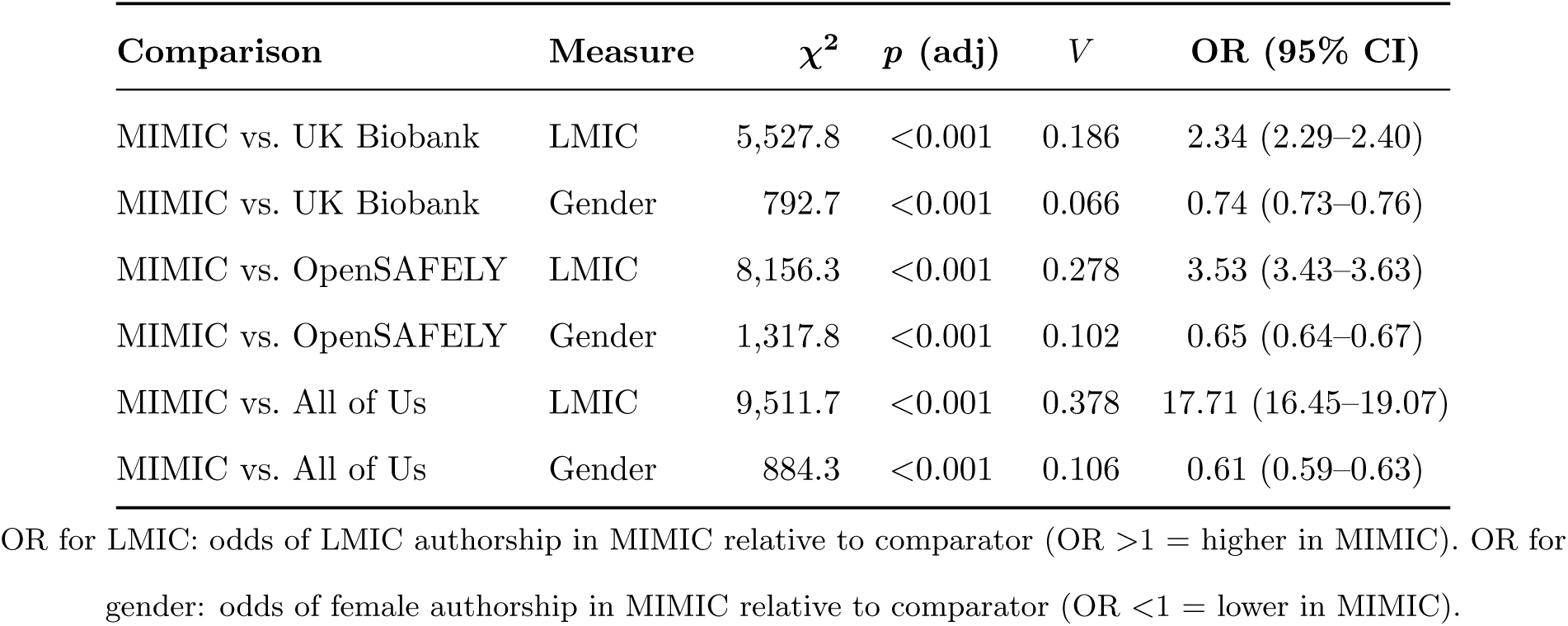
Pairwise chi-square tests comparing MIMIC with each other repository for LMIC representation and gender distribution. All *p*-values are Bonferroni-corrected (*n* = 3).

**Table S2:**
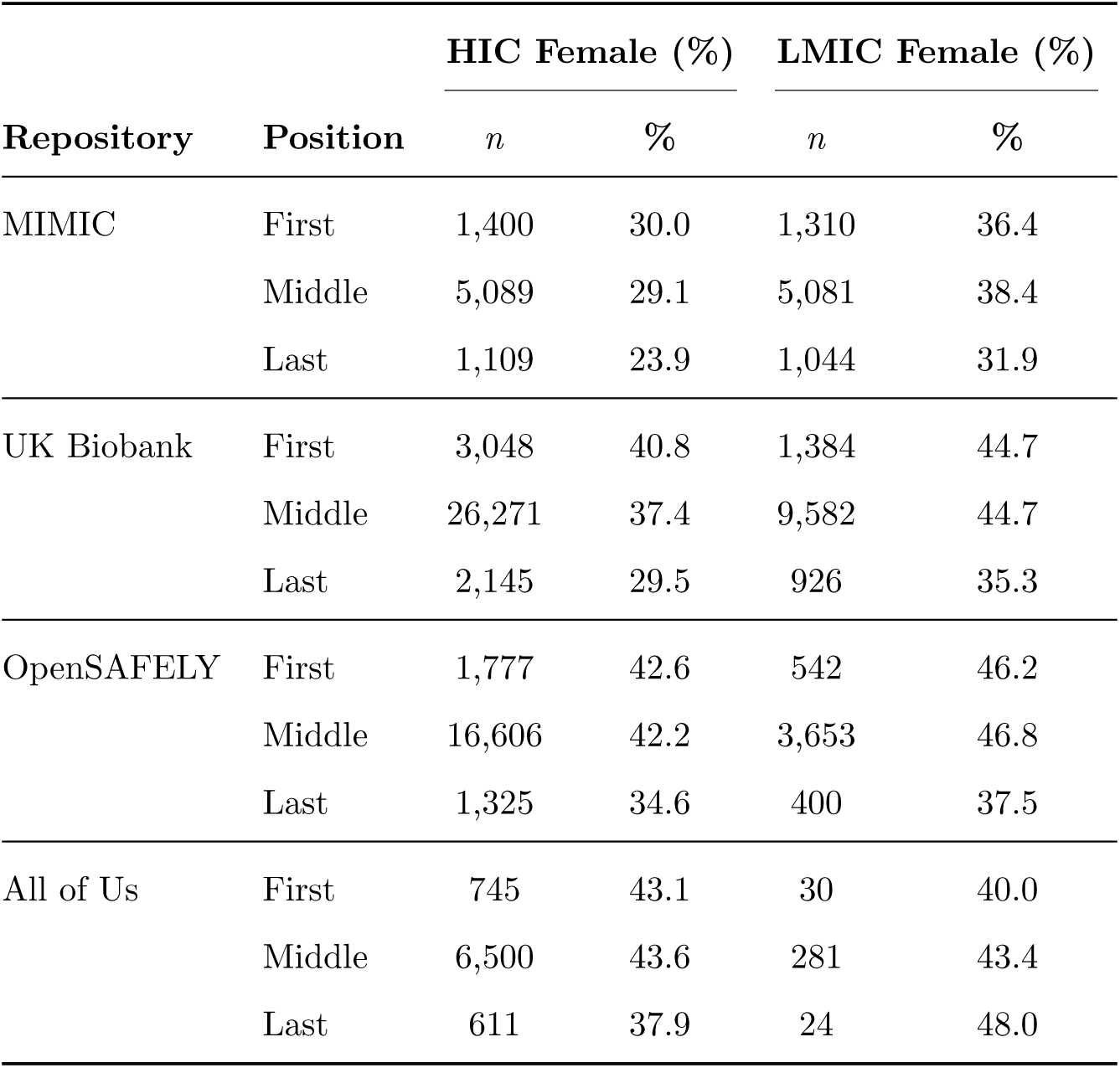
Proportion of female authors within HIC and LMIC subgroups, by authorship position. Female representation is higher among LMIC authors than HIC authors in three of the four repositories (MIMIC, UK Biobank, OpenSAFELY); All of Us shows mixed results with small LMIC sample sizes.

| Repository | Position | HIC Female (%) |  | LMIC Female (%) |  |
| --- | --- | --- | --- | --- | --- |
|  |  | *n* | % | *n* | % |
| MIMIC | First | 1,400 | 30.0 | 1,310 | 36.4 |
|  | Middle | 5,089 | 29.1 | 5,081 | 38.4 |
|  | Last | 1,109 | 23.9 | 1,044 | 31.9 |
| UK Biobank | First | 3,048 | 40.8 | 1,384 | 44.7 |
|  | Middle | 26,271 | 37.4 | 9,582 | 44.7 |
|  | Last | 2,145 | 29.5 | 926 | 35.3 |
| OpenSAFELY | First | 1,777 | 42.6 | 542 | 46.2 |
|  | Middle | 16,606 | 42.2 | 3,653 | 46.8 |
|  | Last | 1,325 | 34.6 | 400 | 37.5 |
| All of Us | First | 745 | 43.1 | 30 | 40.0 |
|  | Middle | 6,500 | 43.6 | 281 | 43.4 |
|  | Last | 611 | 37.9 | 24 | 48.0 |

**Table S3:** Unique author demographics by repository. Authors are deduplicated by OpenAlex author ID within each dataset.

|  | MIMIC | UK Biobank | OpenSAFELY | All of Us |
| --- | --- | --- | --- | --- |
| Unique authors | 33,573 | 51,783 | 44,441 | 13,238 |
| Female (%) | 33.5 | 41.3 | 43.7 | 44.4 |
| LMIC (%) | 47.3 | 30.2 | 21.8 | 5.2 |
| Ever first author ($n$) | 6,921 | 7,278 | 4,810 | 1,517 |
| Female (%) | 32.8 | 42.7 | 43.0 | 43.8 |
| LMIC (%) | 39.2 | 31.3 | 21.3 | 3.8 |
| Ever last author ($n$) | 5,853 | 5,071 | 4,289 | 1,271 |
| Female (%) | 28.3 | 32.8 | 35.5 | 39.4 |
| LMIC (%) | 40.7 | 27.0 | 21.5 | 3.3 |

“Ever first/last author” = unique authors who held that position on at least one publication citing the repository.
MIMIC’s unique LMIC authorship (47.3%) exceeds its record-level rate (41.8%) because LMIC authors appear on fewer papers on average.

**Table S4:** Genderize.io gender identification rates (%) by World Bank region and repository. Rates represent the proportion of unique authors for whom Genderize.io returned a gender assignment. South Asia consistently shows the lowest identification rates across repositories.

| Region | MIMIC | UK Biobank | OpenSAFELY | All of Us |
| --- | --- | --- | --- | --- |
| | $n$ (%) | $n$ (%) | $n$ (%) | $n$ (%) |
| North America | 8,321 (99.2) | 11,975 (99.2) | 7,772 (99.4) | 8,320 (99.2) |
| Europe & Central Asia | 5,935 (98.9) | 16,304 (99.1) | 18,300 (98.4) | 1,920 (99.5) |
| East Asia & Pacific | 15,135 (98.7) | 16,571 (98.9) | 5,362 (98.6) | 1,124 (99.4) |
| Middle East & North Africa | 741 (99.1) | 449 (99.6) | 2,002 (99.5) | 99 (100.0) |
| Latin America & Caribbean | 492 (99.4) | 377 (98.9) | 2,234 (99.3) | 163 (99.4) |
| South Asia | 1,019 (96.8) | 318 (97.8) | 1,084 (97.0) | 71 (95.8) |
| Sub-Saharan Africa | 92 (97.8) | 138 (98.6) | 744 (98.8) | 103 (99.0) |
| Unknown/Unclassified | 4,881 (98.2) | 5,651 (98.6) | 6,943 (97.5) | 1,438 (99.0) |
| Total | 36,616 (98.7) | 51,783 (99.0) | 44,441 (98.5) | 13,238 (99.2) |

Identification rates for South Asia (95.8–97.8%) are systematically lower than other regions, consistent with known limitations of name-based gender inference for South and Southeast Asian names [23].

## References

[1] National Institutes of Health. NIH data sharing policy and implementation guidance, 2023. URL https://sharing.nih.gov/data-management-and-sharing-policy. Accessed: 2026-02-01.

[2] Wellcome Trust. Open research at Wellcome, 2017. URL https://wellcome.org/what-we-do/our-work/open-research. Accessed: 2026-02-01.

[3] Jennifer C. Molloy. The open knowledge foundation: open data means better science. PLoS biology, 9(12):e1001195, 2011. ISSN 1545-7885. doi: 10.1371/journal.pbio.1001195.

[4] Mark D. Wilkinson, Michel Dumontier, I. Jsbrand Jan Aalbersberg, Gabrielle Appleton, Myles Axton, Arie Baak, Niklas Blomberg, Jan-Willem Boiten, Luiz Bonino da Silva Santos, Philip E. Bourne, Jildau Bouwman, Anthony J. Brookes, Tim Clark, Mercè Crosas, Ingrid Dillo, Olivier Dumon, Scott Edmunds, Chris T. Evelo, Richard Finkers, Alejandra Gonzalez-Beltran, Alasdair J. G. Gray, Paul Groth, Carole Goble, Jeffrey S. Grethe, Jaap Heringa, Peter A. C. ’t Hoen, Rob Hooft, Tobias Kuhn, Ruben Kok, Joost Kok, Scott J. Lusher, Maryann E. Martone, Albert Mons, Abel L. Packer, Bengt Persson, Philippe Rocca-Serra, Marco Roos, Rene van Schaik, Susanna-Assunta Sansone, Erik Schultes, Thierry Sengstag, Ted Slater, George Strawn, Morris A. Swertz, Mark Thompson, Johan van der Lei, Erik van Mulligen, Jan Velterop, Andra Waagmeester, Peter Wittenburg, Katherine Wolstencroft, Jun Zhao, and Barend Mons. The FAIR guiding principles for scientific data management and stewardship. Scientific Data, 3:160018, 2016. ISSN 2052-4463. doi: 10.1038/sdata.2016.18.

[5] All of Us Research Program Investigators, Joshua C. Denny, Joni L. Rutter, David B. Goldstein, Anthony Philippakis, Jordan W. Smoller, Gwynne Jenkins, and Eric Dishman. The ”all of us” research program. The New England Journal of Medicine, 381(7):668–676, 2019. ISSN 1533-4406. doi: 10.1056/NEJMsr1809937.

[6] Cathie Sudlow, John Gallacher, Naomi Allen, Valerie Beral, Paul Burton, John Danesh, Paul Downey, Paul Elliott, Jane Green, Martin Landray, Bette Liu, Paul Matthews, Giok Ong, Jill Pell, Alan Silman, Alan Young, Tim Sprosen, Tim Peakman, and Rory Collins. UK biobank: an open access resource for identifying the causes of a wide range of complex diseases of middle and old age. PLoS medicine, 12(3):e1001779, 2015. ISSN 1549-1676. doi: 10.1371/journal.pmed.1001779.

[7] Alistair E. W. Johnson, Tom J. Pollard, Lu Shen, Li-Wei H. Lehman, Mengling Feng, Mohammad Ghassemi, Benjamin Moody, Peter Szolovits, Leo Anthony Celi, and Roger G. Mark. MIMIC-III, a freely accessible critical care database. Scientific Data, 3:160035, 2016. ISSN 2052-4463. doi: 10.1038/sdata.2016.35.

[8] E. Garfield. Citation indexes for science; a new dimension in documentation through association of ideas. Science, 122(3159):108–111, 1955. ISSN 0036-8075. doi: 10.1126/science.122.3159.108.

[9] H. F. Moed. *Citation analysis in research evaluation*. Number v. 9 in Information science and knowledge management. Springer, 2005. ISBN 978-1-4020-3713-9 978-1-4020-3714-6.

[10] Vincent Larivière, Chaoqun Ni, Yves Gingras, Blaise Cronin, and Cassidy R. Sugimoto. Bibliometrics: global gender disparities in science. Nature, 504(7479):211–213, 2013. ISSN 1476-4687. doi: 10.1038/504211a.

[11] Leo Anthony Celi, Jacqueline Cellini, Marie-Laure Charpignon, Edward Christopher Dee, Franck Dernoncourt, Rene Eber, William Greig Mitchell, Lama Moukheiber, Julian Schirmer, Julia Situ, Joseph Paguio, Joel Park, Judy Gichoya Wawira, Seth Yao, and for MIT Critical Data. Sources of bias in artificial intelligence that perpetuate healthcare disparities-a global review. PLOS digital health, 1(3):e0000022, 2022. ISSN 2767-3170. doi: 10.1371/journal.pdig.0000022.

[12] Nick Couldry and Ulises Ali Mejias. *The costs of connection: how data is colonizing human life and appropriating it for capitalism*. Culture and economic life. Stanford University Press, 2019. ISBN 978-1-5036-0366-0 978-1-5036-0974-7.

[13] Miranda Fricker. Epistemic injustice: power and the ethics of knowing. Oxford University Press, repr edition, 2011. ISBN 978-0-19-957052-2.

[14] Alistair E. W. Johnson, Lucas Bulgarelli, Lu Shen, Alvin Gayles, Ayad Shammout, Steven Horng, Tom J. Pollard, Sicheng Hao, Benjamin Moody, Brian Gow, Li-Wei H. Lehman, Leo A. Celi, and Roger G. Mark. MIMIC-IV, a freely accessible electronic health record dataset. Scientific Data, 10(1):1, 2023. ISSN 2052-4463. doi: 10.1038/s41597-022-01899-x.

[15] Clare Bycroft, Colin Freeman, Desislava Petkova, Gavin Band, Lloyd T. Elliott, Kevin Sharp, Allan Motyer, Damjan Vukcevic, Olivier Delaneau, Jared O’Connell, Adrian Cortes, Samantha Welsh, Alan Young, Mark Effingham, Gil McVean, Stephen Leslie, Naomi Allen, Peter Donnelly, and Jonathan Marchini. The UK biobank resource with deep phenotyping and genomic data. Nature, 562(7726):203–209, 2018. ISSN 1476-4687. doi: 10.1038/s41586-018-0579-z.

[16] Elizabeth J. Williamson, Alex J. Walker, Krishnan Bhaskaran, Seb Bacon, Chris Bates, Caroline E. Morton, Helen J. Curtis, Amir Mehrkar, David Evans, Peter Inglesby, Jonathan Cockburn, Helen I. McDonald, Brian MacKenna, Laurie Tomlinson, Ian J. Douglas, Christopher T. Rentsch, Rohini Mathur, Angel Y. S. Wong, Richard Grieve, David Harrison, Harriet Forbes, Anna Schultze, Richard Croker, John Parry, Frank Hester, Sam Harper, Rafael Perera, Stephen J. W. Evans, Liam Smeeth, and Ben Goldacre. Factors associated with COVID-19-related death using OpenSAFELY. Nature, 584(7821):430–436, 2020. ISSN 1476-4687. doi: 10.1038/s41586-020-2521-4.

[17] Andrea H. Ramirez, Lina Sulieman, David J. Schlueter, Alese Halvorson, Jun Qian, Francis Ratsimbazafy, Roxana Loperena, Kelsey Mayo, Melissa Basford, Nicole Deflaux, Karthik N. Muthuraman, Karthik Natarajan, Abel Kho, Hua Xu, Consuelo Wilkins, Hoda Anton-Culver, Eric Boerwinkle, Mine Cicek, Cheryl R. Clark, Elizabeth Cohn, Lucila Ohno-Machado, Sheri D. Schully, Brian K. Ahmedani, Maria Argos, Robert M. Cronin, Christopher O’Donnell, Mona Fouad, David B. Goldstein, Philip Greenland, Scott J. Hebbring, Elizabeth W. Karlson, Parinda Khatri, Bruce Korf, Jordan W. Smoller, Stephen Sodeke, John Wilbanks, Justin Hentges, Stephen Mockrin, Christopher Lunt, Stephanie A. Devaney, Kelly Gebo, Joshua C. Denny, Robert J. Carroll, David Glazer, Paul A. Harris, George Hripcsak, Anthony Philippakis, Dan M. Roden, and All of Us Research Program. The all of us research program: Data quality, utility, and diversity. Patterns, 3(8):100570, 2022. ISSN 2666-3899. doi: 10.1016/j.patter.2022.100570.

[18] George B. Moody and Roger G. Mark. A database to support development and evaluation of intelligent intensive care monitoring. Computers in Cardiology 1996, pages 657–660, 1996. URL https://api.semanticscholar.org/CorpusID:28498264.

[19] Ben Goldacre and Jessica Morley. Better, broader, safer: using health data for research and analysis. Technical report, UK Department of Health and Social Care, 2022.

[20] Jason Priem, Heather Piwowar, and Richard Orr. OpenAlex: A fully-open index of scholarly works, authors, venues, institutions, and concepts, 2022. URL https://arxiv.org/abs/2205.01833. Version Number: 2.

[21] Genderize.io. Genderize.io: Determine the gender of a name, 2024. URL https://genderize.io. Accessed: 2026-02-01.

[22] World Bank. World bank country and lending groups, 2024. URL https://datahelpdesk.worldbank.org/knowledgebase/articles/906519. Accessed: 2026-02-01.

[23] Lucía Santamaría and Helena Mihaljević. Comparison and benchmark of name-to-gender inference services. PeerJ. Computer Science, 4:e156, 2018. ISSN 2376-5992. doi: 10.7717/ peerj-cs.156.

[24] Marc J. Lerchenmueller, Olav Sorenson, and Anupam B. Jena. Gender differences in how scientists present the importance of their research: observational study. BMJ, 367:l6573, 2019. ISSN 1756-1833. doi: 10.1136/bmj.l6573.

[25] Cassidy R. Sugimoto and Vincent Larivière. Equity for women in science: dismantling systemic barriers to advancement. Harvard University Press, 2023. ISBN 978-0-674-91929-7.

[26] Mariam Asghar, Muhammad Shariq Usman, Rafi Aibani, Hamza Tahir Ansari, Tariq Jamal Siddiqi, Kaneez Fatima, Muhammad Shahzeb Khan, and Vincent M. Figueredo. Sex differences in authorship of academic cardiology literature over the last 2 decades. Journal of the American College of Cardiology, 72(6):681–685, 2018. ISSN 1558-3597. doi: 10.1016/j.jacc.2018.05.047.

[27] Milit S. Patel, Maria Jocelyn Kara Magsanoc-Alikpala, Erin Jay G. Feliciano, Leo Anthony Celi, T. Peter Kingham, Nancy Y. Lee, and Edward Christopher Dee. Integrating epistemic justice in global cancer research. Nature Health, 2026. ISSN 3005-0693. doi: 10.1038/s44360-025-00047-0. URL https://www.nature.com/articles/s44360-025-00047-0.

[28] Maui Hudson, Stephanie Russo Carroll, Jane Anderson, Darrah Blackwater, Felina M. Cordova-Marks, Jewel Cummins, Dominique David-Chavez, Adam Fernandez, Ibrahim Garba, Danielle Hiraldo, Mary Beth Jäger, Lydia L. Jennings, Andrew Martinez, Rogena Sterling, Jennifer D. Walker, and Robyn K. Rowe. Indigenous peoples’ rights in data: a contribution toward indigenous research sovereignty. Frontiers in Research Metrics and Analytics, 8:1173805, 2023. ISSN 2504-0537. doi: 10.3389/frma.2023.1173805.

